# A Text Message Intervention to Support Latino Dementia Family Caregivers (CuidaTEXT): Feasibility study

**DOI:** 10.1101/2022.04.12.22273809

**Authors:** Jaime Perales-Puchalt, Mariana Ramírez-Mantilla, Mónica Fracachán-Cabrera, Eric D. Vidoni, Edward F. Ellerbeck, A. Susana Ramírez, Amber Watts, Kristine Williams, Jeffrey Burns

## Abstract

**Objectives:** To test the feasibility, acceptability, and preliminary efficacy of *CuidaTEXT:* a tailored text message intervention to support Latino dementia family caregivers.

**Methods:** *CuidaTEXT* is a six-month, bilingual, and bidirectional intervention tailored to caregiver needs (e.g., education, problem-solving, resources). We enrolled 24 Latino caregivers in a one-arm trial, and assessed feasibility, acceptability, and preliminary efficacy within six months.

**Results:** Recruitment took 61 days and enrollment took 20. None of the participants unsubscribed from *CuidaTEXT*, and 83.3% completed the follow up survey. Most participants (85.7%) reported reading most text messages thoroughly. Participants reported high levels of satisfaction with the intervention (3.6 on a scale from 1 to 4). *CuidaTEXT* helpfulness was high (3.5-3.8 on a 1 to 4 scale). Compared to baseline, at six months caregiver behavioral symptom distress (0–60) decreased from 19.8 to 12.0, and depression (0-30) from 8.8 to 5.4 (p<0.05).

**Conclusions:** *CuidaTEXT* demonstrated high levels of feasibility, acceptability, and preliminary efficacy among Latino caregivers.

**Clinical implications:** *CuidaTEXT’s* feasibility and potential for widespread implementation holds promise in supporting Latino caregivers of people with dementia.

## Introduction

Support for family caregivers of individuals with dementia (IWDs) is a key component of the US National Alzheimer’s Project Act (U.S. Department of Health & Human Services, 2016). Latino caregivers’ physical and mental health is disproportionally impacted by caregiving (Gallagher-Thompson et al., 2003; Garcia, 2000; Hinton et al., 2006; Hinton et al., 2003; Napoles et al., 2010; National Alliance for Caregiving, 2008; Pinquart & Sörensen, 2005; Talamantes & Aranda, 2004), and despite their high interest in receiving caregiver support (Perales et al., 2018), Latinos experience disparities accessing it (Monahan et al., 1992; Scharlach et al., 2008). Family caregiver support interventions have shown efficacy in improving health outcomes (Walter & Pinquart, 2020), but most have been designed for non-Latino Whites and results usually do not generalize to other groups potentially due to linguistic, cultural, and contextual reasons (Gilmore-Bykovskyi et al., 2018; Gitlin et al., 2015; Pendergrass et al., 2015). There is a crucial need for targeted caregiver support interventions among Latinos. This need is in line with the National Institute on Aging’s call to address health disparities in aging research (National Institute on Aging, 2018).

To address Latinos’ caregiving disparities, we developed *CuidaTEXT* (a Spanish play on words for self-care and texting) (Perales-Puchalt, Acosta-Rullan, et al., 2021). *CuidaTEXT* is to our knowledge, the first text message intervention for caregiver support of individuals with dementia (IWDs) among Latinos or any other ethnic group. Text messaging is a well-suited modality to deliver caregiver support for Latinos given its universal use, and engagement in this population, its convenience, low cost, privacy, and scalability (Cartujano-Barrera et al., 2020; Guerriero et al., 2013; Hall et al., 2015; Pew Research Center, 2021; Schilling et al., 2013; Zurovac et al., 2012). The potential of text message interventions among Latinos contrasts with synchronous interventions, or interventions that largely rely on apps, computers or internet broadband, as these may widen disparities among Latinos due to disproportionately lower access (Atske & Perrin, 2021; Katz et al., 2022). The present study aimed to test the feasibility, acceptability, and preliminary efficacy of *CuidaTEXT* among Latino family caregivers of IWDs. This development corresponds to Stage 1b of the NIH Stage Model for Behavioral Intervention Development (feasibility testing) (Onken et al., 2014).

## Methods

This study used a one-arm pre- post-intervention trial design with assessments conducted at baseline and six months. We recruited caregiver participants from June to August 2021 from our center’s clinic, research registries formed in the past five years (Perales-Puchalt, 2020; Perales-Puchalt et al., 2020; Perales et al., 2018) community promotion (newspaper ads, presentations), and advertisements in national organization registries and websites. All participants were enrolled over a 20-day period during the month of August 2021. Participants were eligible if they spoke Spanish or English, were 18 or older, identified as Latino, owned a cellphone with a flat fee, and reported being able to read and write. To be eligible, participants also had to provide hands-on care for a relative with a clinical or research dementia diagnosis, who also scored two or higher in the AD8 proxy-administered cognitive screener (Galvin et al., 2005; Pardo et al., 2013). In our previous research, advisory board members highlighted the importance of allowing the inclusion of more than one caregiver per IWD to reduce burden and increase social support (Perales-Puchalt, Acosta-Rullan, et al., 2021). For this reason, we allowed more than one caregiver participant per IWD. All study procedures were approved by the Institutional Review Board of the University of Kansas Medical Center (STUDY00144478). All participants gave written informed consent.

### Procedures

The research team explained the general characteristics of the study to potential participants over the phone or via secure videoconference. Those willing to participate were screened for eligibility. If eligible, the research team asked caregiver participants to sign an online informed consent and scheduled a phone call or videoconference to complete the baseline assessment. All participants who completed the baseline assessments were considered, enrolled in the study, and immediately began to receive *CuidaTEXT’s* text messages. Six months after the baseline assessments, the research team messaged participants notifying them of their intention to call them and schedule the follow-up assessment, which took place within a two-week window.

### Intervention

*CuidaTEXT* is a bilingual, bidirectional, six-month intervention tailored to caregiver needs via Short Message Service text messages. An in-depth description of the intervention and its development has been previously reported (Perales-Puchalt, Acosta-Rullan, et al., 2021). The intervention is informed by the Stress Process Framework and Social Cognitive Theory (Bandura, 1972; Pearlin et al., 1990). These messages include the identification of barriers to desired behaviors (e.g., problem solving, relaxation techniques, or exercising), setting of realistic goals, encouragement of gradual practice to increase healthy behaviors, integration of testimonials and videos to promote vicarious learning, integration of praise provide social support, and education to increase dementia knowledge. *CuidaTEXT* includes: 1) 1-3 daily automatic messages (n=244 over six months) about logistics, dementia education, self-care, social support, end-of-life care, care of the person with dementia, behavioral symptoms and problem-solving strategies; 2) up to 783 keyword-driven text messages providing on-demand help for the above topics; 3) live chat interaction with a coach from the research team for further help upon request; 4) a 19-page reference booklet summarizing the purpose and functions of the intervention.

### Assessment

The research team collected information from three sources: baseline survey, six-month follow-up survey, and metrics of text message interactions. Pre-intervention survey socio-demographic information included the caregivers’ age, gender, race, US region of residence, and marital and medical insurance status. Acculturation information included the caregiver’s country of birth and primary language (Spanish, English, both, and other). Technological information included whether caregivers had previously registered in another text message notification service (e.g., bank or clinic notifications). Caregiving characteristics included the caregiver’s relationship to the IWD. Care recipients’ characteristics included the IWD’s age, gender, ethnicity, diagnosis, medical insurance status and AD8 cognitive screening score (Galvin et al., 2005; Pardo et al., 2013).

Outcomes included feasibility, acceptability, and preliminary efficacy:

1. <u>Feasibility outcomes</u> included the duration of recruitment and enrollment (study recruitment and enrollment feasibility), percentage of potential participants who opted into *CuidaTEXT* (intervention enrollment feasibility), percentage of participants who completed the follow-up survey (retention feasibility), percentage of enrolled participants who completed all outcome assessments (assessment feasibility), average number of text messages sent by participants, percentage of participants who unsubscribed from *CuidaTEXT* by texting the keyword STOP, percentage of participants who sent at least one text message, and percentage of participants who reported ‘I read through the text messages thoroughly most times’ in the follow-up survey as opposed to ‘I took only a short look at the text messages most times’, or ‘I did not read the text messages most times’ (engagement feasibility). The follow-up survey also included a free-response question asking whether participants experienced any technical problems (intervention delivery feasibility).
2. <u>Acceptability outcomes</u> were all collected in the follow-up survey. These outcomes included nine four-point Likert scale questions on satisfaction with *CuidaTEXT* and its components (Not at All-Extremely). Three additional four-point Likert scale questions asked about their perceived helpfulness of *CuidaTEXT* in: caring for the IWD, caring for themselves, and enhancing their understanding of dementia (Not at All-A Lot). Each question had a slot for comments, which the interviewee typed in describing them. The survey also included the System Usability Scale, which asks about their experience with the *CuidaTEXT* (Sauro, 2011). The System Usability Scale is a valid and reliable 10-item questionnaire Likert scale (1-5). According to the developers of the scale, scores above 68 out of 100 indicate higher levels of usability.
3. <u>Preliminary efficacy outcomes</u> included scales administered at baseline and follow-up. Most of these scales were validated in the US Latino Spanish-speaking population. For those that were not, we used either Spanish-speaking versions from other countries, or we translated them using standard procedures (World Health Organization, 2018). The first two outcomes include proxy scales that refer to the IWD and all the others refer to the caregiver:

Functional Activities Questionnaire (FAQ) (Acevedo et al., 2009; Pfeffer et al., 1982). This is a 10-item questionnaire completed by the caregiver as a proxy respondent for the care recipient to monitor changes in instrumental activities of daily living over time (e.g., preparing balanced meals, following the news, or playing a game of skill). Each item is rated with six response options: Dependent (3 points), Requires Assistance (2 points), Has Difficulty but Does By Self (1 point), Normal (0 points), Never Did but Could Do Now (0 points), and Never Did And Would Have Difficulty Now (1 point). Total scores range from 0 to 30 with higher scores indicating higher dependence. Given the nature of dementia, this outcome is expected to either remain stable or to worsen over time, but the information may be helpful to compare how other outcomes change despite the functional decline.

Behavioral symptom severity (NPI-Q) (Acevedo et al., 2009; Kaufer et al., 2000). The NPI-Q is a clinical instrument for evaluating psychopathology in dementia with two scales, care recipient severity (NPI-Q-S) and caregiver distress (NPI-Q-D). If any of the 12 neuropsychiatric symptoms is present in the last month (e.g., depression, repeating, delusions), caregivers rate the level of severity for the IWDs on a 3-point scale (Mild–Severe). An overall severity summary score is calculated by adding the severity scores of all items. For any present symptom, caregivers also rate their own distress on a 6-point scale (Not Distressing at All-Extreme or Very Severe Distress). An overall distress summary score is calculated by adding the distress scores of all items. Higher scores indicate higher severity and distress.

Caregiver strain (Modified Caregiver Strain Index) (Thornton & Travis, 2003). The Caregiver Strain Index is a 13-item screener that measures caregiver strain. For each of the items, the caregiver can respond either No (0), Yes Sometimes (1), or Yes on a Regular Basis (2). The total score is the sum of all item scores. Higher scores indicate higher strain.

Caregiver burden (Zarit Burden Interview-6; ZBI-6) (Higginson et al., 2010). The ZBI-6 measures caregiver burden. Each of the six items of the ZBI-6 is a statement the caregiver is asked to endorse using a 5-point scale. Response options range from 0 (Never) to 4 (Nearly Always). An overall burden summary score is calculated by adding the scores of all items, and higher scores indicate higher burden.

Caregiving competence (Preparedness for Caregiving Scale; PCS) (Carter et al., 1998; Gutierrez-Baena & Romero-Grimaldi, 2021). The PCS is a self-rated instrument that consists of eight items that ask caregivers how well prepared they believe they are for multiple domains of caregiving. Responses are rated on a 5-point Likert scale with scores ranging from 0 (Not at All Prepared) to 4 (Very Well Prepared). The scale is scored by calculating the mean of all items answered with a total score range of 0 to 4. The higher the score the more prepared the caregiver feels for caregiving.

Positive aspects of caregiving (Positive aspects of caregiving scale) (Tarlow et al., 2004). This scale was designed to measure psychosocial benefits of caregiving among family caregivers. The scale has nine statements and caregivers rate their level of agreement with those statements ranging from 1 (Disagree A Lot) to 5 (Agree A Lot). An overall score can be obtained by adding their item scores. Higher scores indicate more perceived positive benefits.

Unmet needs (Measure of Unmet Needs) (Gaugler et al., 2004). The Measure of Unmet Needs is a 24-item survey for caregivers that requires 0 (No) or 1 (Yes) answers regarding additional assistance with categories such as activities of daily living, dementia symptoms, and social support. A total unmet needs score can be obtained by summing all 34 items. The higher the score, the higher the number of unmet needs the caregiver reports having.

ADRD knowledge (Epidemiology/Etiology Disease Scale; EEDS) (Connell & Holmes, 1996; Roberts & Connell, 2000). The EEDS is a 14-item True/False questionnaire about dementia. An example of a question includes ‘There is no cure for Alzheimer’s disease (True)’. Correct answers are scored one point each and a total score is calculated by adding the 14 items. The higher the score, the higher the dementia knowledge.

Social support (Interpersonal Support Evaluation List; ISEL-12) (Merz et al., 2014). The ISEL-12 is a 12-item measure of perceptions of social support. This questionnaire has three different subscales designed to measure dimensions of perceived social support: Appraisal support, belonging support, and tangible support. Each dimension is measured by 4 items on a 4-point scale ranging from ‘Definitely True’ to ‘Definitely False’. Higher scores indicate higher perceived support.

Coping (Coping Orientation to Problems Experienced Inventory; COPE-28) (Carver, 1997; Perczek et al., 2000). The COPE-28 is a 28 item self-report questionnaire designed to measure effective and ineffective ways to cope with caregiving. The scale is rated using a 4-point Likert scale ranging from 1 (I Haven’t Been Doing This at All) to 4 (I Have Been Doing This A Lot). Subscales include problem-focused coping, emotion-focused coping, and avoidant coping. High scores indicate caregiver participants use that strategy more often. Higher scores in problem-focused coping and lower avoidant coping are typically indicative of positive outcomes.

Depressive symptoms (Center for Epidemiologic Studies Depression Scale; CES-D-10) (Cheng & Chan, 2005; González et al., 2017). This is a 10-item, self-report rating scale that measures characteristic symptoms of depression in the past week (e.g., depression, loneliness, restless sleep). Each item is rated on a 4-point scale, from 0 (Rarely or None of the Time) to 3 (Most or All of the Time) with positively worded items (items 5 and 8) reverse scored. Items yield summary scores that range from 0 to 30, with higher scores indicating higher depression severity.

Affect (Scale of Positive and Negative Experience; SPANE) (Daniel-González et al., 2020; Diener et al., 2010). The SPANE comprises 12 items, six positive (SPANE-P) and six negative experiences (SPANE-N). Both sets of items measure three general and three specific emotions encompassing a wide range of human experiences. The instrument uses a five-point frequency rating scale ranging from 1 (Very Rarely or Never) to 5 (Very Often or Always). Total scores range from 6 to 30 with high scores indicating high positive (SPANE-P) or high negative affect (SPANE-N).

Self-perceived health (Patel et al., 2003). This is a one-item question that is self-reported by the caregiver. The question includes a four-point Likert scale and asks ‘Overall, how would you rate your health-Excellent, Good, Fair, or Poor?’.

### Analysis

We used descriptive statistics to summarize baseline characteristics of caregivers and IWDs. We also used descriptive statistics to summarize quantitative feasibility and acceptability outcomes. We summarized acceptability comments and reported the most frequent ones. Regarding preliminary efficacy, we used paired-samples t-tests to assess change from pre-to post-intervention, as all scores were normally distributed. To explore potential mechanisms, we calculated Pearson correlations to analyze between-outcome associations among those outcomes that changed statistically. We used SPSS v20.0 for all calculations (IBM Corp., 2013). The significance level was set at p < 0.05.

## Results

We screened 31 potential caregiver participants. Among those, 24 participants caring for 21 IWDs were enrolled in the study. The reasons for screen failure included no longer being able to participate (n=6) and a lack of research or clinical diagnosis of the IWD (n=1). Participants were recruited from a memory clinic (n=5), research registries (n=6), community promotion (n=5), and advertisements in national registries of caregivers and an organization’s website (n=8). Of the 21 IWDs, 19 had one participating caregiver, one had two caregivers participating and one had three caregivers participating. Given that most IWDs had one participating caregiver, we report the findings of all participants individually. Ancillary analyses with only one participant per IWD (the first one to be enrolled) show similar results (Appendix 1).

Table 1 shows the participants’ characteristics at baseline for the total sample, as well as those who completed the follow-up survey and those who did not. Fourteen participants (58.4%) were caregivers of care recipients with late onset Alzheimer’s dementia, five with dementia in of unspecified etiology (20.8%), three with vascular dementia (12.6%), one with early onset Alzheimer’s dementia, and one with early onset Alzheimer’s dementia and frontotemporal dementia. Caregiver participants’ mean age was 52.6 years and ranged from 26 to 81. Twenty (83.3%) were women and 13 (54.2%) were married or had a partner. Ten participants were born in the USA (41.7%), six in Mexico (25.0%) and eight (33.3%) in another Latin American country. Those not born in the USA had been in the USA for an average of 23.2 years, ranging from 3.0 to 50.0. Eleven (45.8%) chose to receive *CuidaTEXT* messages in Spanish. Most participants (n=18; 75.0%) were the adult children of an IWD.

**Table 1.** Baseline characteristics of the participants enrolled in *CuidaTEXT*.

|  | Total (n=24) | Completers<br>(n=21) | Non-completers<br>(n=3) | P Value |
| --- | --- | --- | --- | --- |
| Age in years, mean (SD) | 52.6 (13.2) | 53.8 (2.3) | 44.0 (15.6) | 0.594 |
| Women, % (n) | 83.3% (20) | 81.0% (17) | 100.0% (3) | 0.563 |
| Region |  |  |  |  |
| Midwest, % (n) | 70.8% (17) | 66.7% (14) | 100.0% (3) | 0.494 |
| South, % (n) | 16.7% (4) | 19.0% (4) | 0.0% (0) |  |
| West, % (n) | 12.5% (3) | 14.3% (3) | 0.0% (0) |  |
| Race, % (n) |  |  |  |  |
| Other/ Hispanic/Latino, % (n) | 62.5% (15) | 61.9% (13) | 66.7% (2) | 0.854 |
| White, % (n) | 29.2% (7) | 28.6% (6) | 33.3% (1) |  |
| Mixed, % (n) | 8.3% (2) | 9.5% (2) | 0.0% (0) |  |
| Country of birth |  |  |  |  |
| US, % (n) | 41.7% (10) | 42.9% (9) | 33.3% (1) | 0.176 |
| Mexico, % (n) | 25.0% (6) | 19.0% (4) | 66.7% (2) |  |
| Other, % (n) | 33.3% (8) | 38.1% (8) | 0.0% (0) |  |
| Years of education, m (SD) | 14.7 (3.8) | 14.9 (3.9) | 13.0 (3.0) | 0.424 |
| Caregiver without medical insurance, % (n) | 20.8% (5) | 23.8% (5) | 0.0% (0) | 0.310 |
| Spanish only as primary language, % (n) | 41.7% (10) | 42.9% (9) | 33.3% (1) | 0.409 |
| Married or have a partner, % (n) | 54.2% (13) | 52.4% (11) | 66.7% (2) | 0.796 |
| Ever signed up for text message reminders, % (n) | 66.7% (16) | 66.7% (14) | 66.7% (2) | 0.723 |
| Relation to care recipient |  |  |  |  |
| Children, % (n) | 75.0% (18) | 76.2% (16) | 66.7% (2) | 0.641 |
| Children-in-law, % (n) | 8.3% (2) | 9.5% (2) | 0.0% (0) |  |
| Partner, % (n) | 16.7% (4) | 14.3% (3) | 33.3% (1) |  |
| Financial inadequacy (0-4), m (SD) | 1.7 (1.3) | 1.7 (0.3) | 1.7 (0.3) | 0.918 |
| Self-rated health (0-4), m (SD) | 2.4 (0.8) | 2.5 (0.2) | 2.0 (0.0) | **0.014** |
| Spirituality (1-5), m (SD) | 4.5 (1.2) | 4.5 (0.3) | 4.7 (0.3) | 0.849 |
| Care recipients is a woman, % (n) * | 61.9% (13) | 66.7% (12) | 33.3% (1) | 0.316 |
| Care recipients is a Latino, % (n) * | 95.2% (20) | 94.4% (17) | 100.0% (3) | 0.857 |
| Age of care recipient, m (SD) * | 74.9 (12.6) | 76.6 (11.7) | 65.0 (15.6) | 0.145 |
| Care recipient without medical insurance, % (n) * | 28.6% (6) | 27.8% (5) | 33.3% (1) | 0.900 |
| AD8 total score of care recipient, m (SD) * | 7.2 (1.3) | 7.5 (0.6) | 7.0 (1.0) | 0.245 |

\* Out of 21 care recipients; **Bold:** p<0.05

Among the 21 IWDs, the average age was 74.9 and ranged from 52.0 to 89.0. Thirteen were women (61.9%), and six (28.6%) had no medical insurance. No baseline caregiver or IWD characteristics were statistically different between those who completed the follow-up assessment and those who did not, except for self-rated health, which was better among completers (mean=2.5) than non-completers (mean=2.0; p=0.014).

Table 2 shows the feasibility and acceptability outcomes.

**Table 2.** Feasibility and acceptability of the *CuidaTEXT* study and intervention (n=21)

|  |  |  |
| --- | --- | --- |
| Feasibility |  |  |
| Study recruitment | Recruitment duration | 61 days |
| Study enrollment | Enrollment duration * | 20 days |
| Intervention enrollment | Percentage of participants able to opt into CuidaTEXT* | 100% (n=24) |
| Retention rate | Percentage of enrolled participants who completed the follow-up survey* | 87.5% (n=21) |
| Assessment feasibility | Percentage of enrolled participants who completed the preliminary efficacy outcomes* | 83.3% (n=20) |
| Intervention delivery | Percentage of participants who experienced technical issues<br>1) Sometimes had no internet to see videos, 2) lost text message history when phone stopped working | 9.5% (n=2) |
| Intervention engagement | Average number of messages sent per participant* | 77.0 (SD=88.4) |
|  | Average number of keywords sent per participant* | 38.0 (SD=69.8) |
|  | Average number of live chat texts sent per participant* | 39.0 (SD=48.4) |
|  | Percentage of participants who sent at least one message* | 91.7% (n=22) |
|  | Percentage of participants who texted STOP to discontinue* | 0% (n=0) |
|  | Percentage of participants who reported reading most messages thoroughly | 85.7% (18) |
| Acceptability |  |  |
| Satisfaction | Overall satisfaction with the intervention (1-4) | 3.6 (SD=0.5) |
|  | Satisfaction with number of texts per day (1-4) | 3.5 (SD=0.7) |
|  | Satisfaction with keyword functionality (1-4) | 3.7 (SD=0.5) |
|  | Satisfaction with links to videos (1-4) | 3.5 (SD=0.8) |
|  | Satisfaction with phone numbers of resources (1-4) | 3.2 (SD=1.0) |
|  | Satisfaction with links to resources (1-4) | 3.7 (SD=0.6) |
|  | Satisfaction with length of the intervention (1-4) | 3.4 (SD=0.9) |
|  | Satisfaction with booklet (1-4) | 3.6 (SD=0.5) |
|  | Satisfaction with family participation (1-4)** | 4.0 (SD=0.0) |
| Helpfulness | To care for their care recipient (1-4) | 3.7 (SD=0.6) |
|  | To care for themselves (1-4) | 3.5 (SD=0.8) |
|  | To understand more about the disease (1-4) | 3.8 (SD=0.7) |
| Usability | System Usability Scale score (0-100) | 95.8 (SD=9.7) |

\* Metrics calculated for all participants (n=24); \*\* only four participants responded to this question, as all other participants enrolled alone with no other relative.

Recruitment was completed in 61 days, and enrollment in 20 days. Among the 24 enrolled caregiver participants, all enrolled (received the initial *CuidaTEXT* message) without any issue, 21 (87.5%) completed at least the acceptability questions of the follow-up survey, and 20 (83.3%) completed the whole survey including the efficacy outcomes. The reason for one participant’s incomplete follow-up survey was due to the death of the participant’s IWD during the intervention (unrelated to their participation). We did not collect the efficacy information for this participant because it would not reflect the effect of the intervention. Among the 24 enrolled participants, none texted the keyword STOP to discontinue the intervention, 22 (91.7%) sent at least one text message to CuidaTEXT and the average number of messages sent to CuidaTEXT was 77.0 (SD=88.4), half of them being keywords and the other half live chat texts (mostly to show gratitude for the messages they received from *CuidaTEXT*). Among the 21 participants who responded to the follow-up survey, 18 reported reading most messages thoroughly (85.7%), and two (9.5%) reported experiencing mild technical issues. One reported sometimes having no internet to see videos that were part of the intervention referred content, and the other reported losing their text message history after they fixed their phone after a technical issue. All the satisfaction and helpfulness outcomes were between ‘Very and Extremely’ satisfactory. For example, the overall satisfaction with *CuidaTEXT* was mean score of 3.6 in a 1 to 4 scale (SD=0.5). All helpfulness outcomes were close to ‘A Lot’. This includes CuidaTEXT’s helpfulness in caring for their loved one with dementia (mean=3.7, SD=0.6), themselves as caregivers (mean=3.5, SD=0.8), and learning about dementia (mean=3.8, SD=0.7). The System Usability Scale mean score was 95.8 (SD=9.7), which is above the high usability threshold of 68.

Caregiver participants’ free-response comments highlighted the helpfulness of *CuidaTEXT* in caring for themselves and their IWD. Participants reported that *CuidaTEXT* improved their dementia understanding/knowledge, perspective/attitudes, skills, and access to resources. *CuidaTEXT* provided a constant feeling of being supported, nudges or reminders for self-care, and validation of their own caregiving actions. The text messaging modality was more manageable than websites and other formats, as it provided daily and on-demand ‘pills of information’. While most participants did not have their relatives enroll in the intervention due to lack of feasibility (e.g., living far away, too busy), some forwarded *CuidaTEXT* content to them. Participants described the content of *CuidaTEXT* as reliable, with diverse, practical, and useful information, with clear language, and easy to access and to digest it.

Table 3 shows the preliminary efficacy of *CuidaTEXT* on caregivers’ assessments of the IWDs and themselves. The caregivers’ ratings of their IWD’s functional dependence as measured by the FAQ did not change from pre- to post-intervention. The overall mean of IWDs’ behavioral symptom severity, as rated by the caregiver in the NPI-Q, decreased from 16.2 to 11.8 (p=0.004). Several caregiver outcomes improved from pre- to post-intervention (p < 0.05): behavioral symptom distress (NPI-Q-D) decreased from 19.8 to 12.0, caregiver strain (CSI) decreased from 13.3 to 10.7, caregiver competence (PCS) increased from 2.1 to 2.6, caregiver unmet needs decreased from 15.7 to 9.4, dementia knowledge (EEDS) increased from 10.0 to 11.2, problem-focused coping (COPE-28) increased from 21.8 to 24.8, depression (CES-D-10) decreased from 8.8 to 5.4, and positive affect (SPANE-P) increased from 23.3 to 25.4. None of the other changes in outcomes were statistically significant, although most followed a trend of improvement.

**Table 3.** Preliminary efficacy outcomes comparing pre- and post-intervention scores.

|  |  | Baseline |  | Follow up |  |  |
| --- | --- | --- | --- | --- | --- | --- |
| **Person with dementia** | N | M | SD | M | SD | P-Value |
| FAQ dependence (0-30) | 20 | 24.3 | 5.0 | 25.5 | 5.3 | 0.237 |
| NPI-Q Severity (0-36) | 20 | 16.2 | 8.0 | 11.8 | 8.9 | **0.004** |
| **Caregiver** |  |  |  |  |  |  |
| NPI-Q Distress (0-60) | 20 | 19.8 | 13.5 | 12.0 | 11.8 | **0.001** |
| Caregiver Strain Index (0-26) | 20 | 13.3 | 5.6 | 10.7 | 6.4 | **0.003** |
| ZBI-6 Burden (0-24) | 20 | 10.4 | 5.4 | 9.1 | 6.2 | 0.179 |
| PCS Competence (0-4) | 20 | 2.1 | 0.7 | 2.6 | 0.7 | **0.000** |
| Positive aspects of caregiving (9-45) | 20 | 33.0 | 7.7 | 34.4 | 7.6 | 0.344 |
| Unmet needs (0-34) | 20 | 15.7 | 8.6 | 9.4 | 6.9 | **0.000** |
| EEDS ADRD knowledge (0-14) | 21 | 10.0 | 1.7 | 11.2 | 1.4 | **0.005** |
| ISEL-12 appraisal support (4-16) | 20 | 12.3 | 3.5 | 13.3 | 3.1 | 0.194 |
| ISEL-12 belonging support (4-16) | 20 | 11.9 | 3.4 | 12.3 | 3.0 | 0.577 |
| ISEL-12 tangible support (4-16) | 20 | 12.5 | 2.5 | 12.9 | 3.0 | 0.587 |
| COPE 28 problem focused (8-32) | 20 | 21.9 | 5.6 | 24.8 | 4.8 | **0.041** |
| COPE 28 emotion focused (12-48) | 20 | 30.9 | 4.9 | 32.7 | 5.8 | 0.120 |
| COPE 28 avoidant focused (8-32) | 20 | 13.1 | 3.4 | 12.3 | 3.1 | 0.234 |
| CES-D-10 depression (0-30) | 20 | 8.8 | 4.3 | 5.4 | 4.1 | **0.008** |
| SPANE-Positive affect (6-30) | 20 | 22.3 | 5.0 | 25.4 | 3.4 | **0.001** |
| SPANE-Negative affect (6-30) | 20 | 13.2 | 3.5 | 11.3 | 3.2 | 0.056 |
| Perceived health (1-4) | 20 | 2.5 | 0.8 | 2.5 | 0.7 | 0.999 |

**Bold:** p<0.05

Table 4 shows the correlations between preliminary efficacy outcomes. The reduction of IWDs’ behavioral symptom severity was only associated with reductions in caregiver behavioral symptom distress (r=0.791). The reduction in caregiver strain was associated with an increase in caregiver competence (r=-0.532) and a reduction in caregiver unmet needs (r=0.457). Increases in caregiver competence were associated with increases in caregiver dementia knowledge (r=0.521) and reductions in depression (r=0.516). In addition to strain, reductions in caregiver unmet needs were associated with increases in dementia knowledge (r=-0.536) and reductions in depression (r=0.735). The only additional statistically significant association was that increases in dementia knowledge were associated with decreases in depression (r=-608; p<0.05 for all reported correlations).

**Table 4.** Mechanistic analysis: between-preliminary efficacy outcome correlations.

|  | NPI-Q<br>Severity<br>change | NPI-Q<br>Distress<br>change | Caregiver<br>Strain<br>Index<br>change | PCS<br>Compete<br>nce<br>change | Unmet<br>needs<br>change | EEDS<br>ADRD<br>knowledg<br>e change | COPE 28<br>problem<br>focused<br>change | CES-D-10<br>depressio<br>n change | SPANE-Positive<br>affect<br>change |
| --- | --- | --- | --- | --- | --- | --- | --- | --- | --- |
| NPI-Q Severity<br>change | 1 | - | - | - | - | - | - | - | - |
| NPI-Q Distress<br>change | **0.791** | 1 | - | - | - | - | - | - | - |
| Caregiver Strain<br>Index change | 0.311 | 0.390 | 1 | - | - | - | - | - | - |
| PCS Competence<br>change | -0.361 | -0.329 | **-0.532** | 1 | - | - | - | - | - |
| Unmet needs<br>change | 0.190 | 0.293 | **0.457** | -0.311 | 1 | - | - | - | - |
| EEDS ADRD<br>knowledge change | -0.123 | -0.051 | -0.373 | **0.521** | **-0.536** | 1 | - | - | - |
| COPE 28 problem<br>focused change | 0.062 | 0.007 | 0.172 | -0.215 | 0.125 | -0.159 | 1 | - | - |
| CES-D-10<br>depression change | 0.321 | 0.240 | 0.281 | **-0.516** | **0.735** | **-0.608** | -0.024 | 1 | - |
| SPANE-Positive<br>affect change | -0.109 | -0.215 | -0.391 | -0.011 | -0.339 | 0.256 | -0.193 | -0.295 | 1 |

**Bold:** p<0.05

## Discussion

This study aimed to test the feasibility, acceptability, and preliminary efficacy of *CuidaTEXT*, a text message intervention to support Latino dementia family caregivers of IWDs. To the best of our knowledge, this is the first caregiver support intervention that relies largely on text messages to deliver its content. This is also one of the few caregiver support interventions purposefully developed to address common linguistic, cultural, and contextual barriers that Latino caregivers experience (Gilmore-Bykovskyi et al., 2018; Gitlin et al., 2015; Pendergrass et al., 2015). We used several survey questions, validated scales, and metrics to assess outcomes during six months among 24 Latino caregivers. Overall, results show that the *CuidaTEXT* study design and intervention are highly feasible, caregiver participants are highly satisfied with the intervention, and the intervention leads to improved outcomes of caregivers and IWDs. These findings are important given that Latino dementia caregivers experience disproportionate levels of physical and mental health issues (Gallagher-Thompson et al., 2003; Garcia, 2000; Hinton et al., 2006; Hinton et al., 2003; Napoles et al., 2010; National Alliance for Caregiving, 2008; Pinquart & Sörensen, 2005; Talamantes & Aranda, 2004), and experience disparities accessing caregiving support (Monahan et al., 1992; Scharlach et al., 2008).

Latinos are rarely included in caregiver support intervention research and are often thought of as ‘hard to reach’ populations (Gilmore-Bykovskyi et al., 2018; Gitlin et al., 2015; Pendergrass et al., 2015). This exclusion raises questions about the generalizability of evidence-based interventions among Latinos, and potentially widens the gap between those for whom interventions were developed and those who were not included (Butler et al., 2020; Gitlin et al., 2015; Pendergrass et al., 2015). Our findings suggest that centering the development of an intervention on Latinos can lead to quick enrollment rates, and high retention rates, usability, levels of intervention engagement, and satisfaction. These results are consistent with *CuidaTEXT’s* user-centered design, which aims to develop usable and acceptable products consists by gathering and incorporating feedback from users into product design (International Organization for Standardization, 2018; Perales-Puchalt, Acosta-Rullan, et al., 2021). In fact, Latinos have been shown to have high interest in receiving caregiver support (Perales et al., 2018). *CuidaTEXT* provides caregivers with remote and asynchronous ways to receive support, increasing their access to support services. Remote assessments also increases the feasibility of the study design, making it even more valuable during the COVID-19 pandemic.

*CuidaTEXT* resulted in decreased levels of caregiver’s perceived behavioral symptom severity among IWDs, and caregivers’ distress, strain, and depression, among other outcomes after six months. Improvements in these outcomes took place during the COVID-19 pandemic, when there is evidence of worsening of these outcomes (Gedde et al., 2022), and for which some tele-psychological support to caregivers (IWD or caregiver-focused) did not show improvements from baseline to 28- and 32-week follow-up assessments (Rotondo et al., 2021). The preliminary efficacy of *CuidaTEXT* is comparable to in-person interventions with the additional benefit that it is less workforce-intensive and can be delivered remotely and almost fully automatically. Our analyses showed a 40.0% decrease in the average number of caregiver unmet caregiver needs, and a 19.5% reduction in feelings of caregiver strain. These results are in line with the effectiveness of the Reducing Disability in Alzheimer’s Disease intervention, one of the few evidence-based interventions that have been implemented in the community (Menne et al., 2014; Perales-Puchalt, Barton, et al., 2021). *CuidaTEXT* led to average behavioral symptom severity score reductions of 4.4 points, and behavioral symptom distress reductions of 7.8 points more than the 2.8 and 3.1 points reported respectively to be considered a minimally clinically significant difference using the same scale (Mao et al., 2015). Other studies using the same depression scale found that control group participants reported average reductions of 0.2, 0.2, and 0.7 after five or six months (Czaja et al., 2013; Finkel et al., 2007; Martindale-Adams et al., 2013). This difference is smaller than the average 3.4 difference found in our study, which is the equivalent of one depressive symptom over 5-7 days, or three depressive symptoms over 1-2 days in the last week. In fact, this reduction in depressive symptoms is comparable or bigger than the reductions reported by caregivers receiving the active caregiver support intervention in those controlled trials (Czaja et al., 2013; Finkel et al., 2007; Martindale-Adams et al., 2013). *CuidaTEXT* also led to increases in positive affect, which is an important outcome that has been rarely included in clinical trials, likely due to the prevailing deficit focus of biomedical research (de Manincor et al., 2016; Espejo et al., 2020).

Both participants’ comments and preliminary efficacy outcomes gave insight into potential mechanisms of the intervention. For example, caregiver participants reported that *CuidaTEXT* helped improve their understanding of dementia, attitudes, skills for caregiving, and access to resources. These comments were corroborated by improvements in dementia knowledge, competence, and problem-related coping, and reductions in caregiver unmet needs. In fact, these outcomes were associated with improvements in caregiver depression and strain, which suggest a potential mechanism. Therefore, *CuidaTEXT* might reduce caregiver depression and strain by improving dementia knowledge and competence, and by reducing their unmet needs. This mechanism is in line with the Stress Process Framework that was used to inform *CuidaTEXT* (Pearlin et al., 1990). Caregivers’ frequent comments about *CuidaTEXT* providing a constant source of support did not have a corresponding increase in the social support scale. However, the ISEL-12 focuses on family and friends, and it likely does not reflect the support participants received from *CuidaTEXT*. Future studies should use a social support scale that better reflects this source of support.

This study has some limitations. The pre-post design with no control group prevents our ability to infer causal relationships between the intervention and observed outcomes. We were unable to maintain contact for follow-up with three participants, who might have had a more negative feedback and outcomes than the participants who completed the follow-up assessments. The small sample size and its convenience sampling may reduce the generalizability of these findings. The average level of functional dependence of IWDs was relatively high, which raises the question of whether *CuidaTEXT* would achieve such strong outcomes among samples with a lower level of dependence. Our sample had a distribution that was similar to the US Latino caregiver population in terms of language and medical insurance status. However, despite men’s low participation in dementia caregiving, their distribution in our study (16%) was smaller than the Latino caregiving population (26%), likely due to Latino men’s previously reported lower participation in research (National Alliance for Caregiving, 2008). Given the nature of the study, neither the participants, nor the assessment staff nor the data analyst were blinded, which could have biased the results. The current study did not analyze the content of text messages sent by caregivers. The content analysis is out of the scope of this manuscript and will be reported in a future manuscript using mixed-methods.

This study has implications for public health, clinical practice, and research. *CuidaTEXT* has high potential for implementation, given the universal accessibility of Short Message Service text messaging and its reliance on technology rather than workforce, which potentially makes it more cost-effective and sustainable. The positive feasibility and acceptability findings in this study highlights the importance for intervention developers to design interventions with implementation in mind from the beginning of the intervention, and using user-centered design for its future success (Gaugler et al., 2021; International Organization for Standardization, 2018). The next logical step for *CuidaTEXT* is an efficacy randomized controlled trial, which corresponds with Stage 2 of the NIH Stage Model for Behavioral Intervention Development (Onken et al., 2014). If successful, *CuidaTEXT* could be easily implemented in clinics and community organizations, by having the caregivers send a text message to enroll or by having staff enter their phone numbers and names on a website. A future Stage 2 trial should also aim to assess mechanisms of action. Our current study suggests some mechanisms that can be tested in that future study. Despite the encouragement from our advisory board to recruit more than one caregiver per IWD due to Latinos’ family distribution of caregiving tasks (Perales-Puchalt, Acosta-Rullan, et al., 2021), most participants decided to participate on their own. Future studies should explore how to increase engagement of other family members, although our participants have already provided a valid solution: forwarding the messages they consider important to their families. Given its many advantages (e.g., on-request tailoring, available anywhere and at any time, low dependence on workforce) future studies could explore the feasibility of *CuidaTEXT* in other populations in the US and elsewhere, including rural areas and low- and middle-income countries.

## Conclusion

This study tested the feasibility, acceptability, and preliminary efficacy of *CuidaTEXT*, the first tailored text message intervention specifically designed to support dementia family caregivers in the Latino community. This study design was feasible in this underserved population, and the intervention showed high levels of usability, engagement, and satisfaction, and a promising increase in important clinical caregiver-reported outcomes. These positive findings and the potential for widespread implementation, support *CuidaTEXT* as an ideal intervention to eliminate caregiver dementia disparities among Latinos, a key aim in US federal policies (National Institute on Aging, 2018; U.S. Department of Health & Human Services. Office of The Assistant Secretary for Planning and Evaluation, 2020).

### Clinical Implications

- Latino family caregivers of individuals with dementia face many barriers to caregiver support access that may be alleviated through culturally tailored text message interventions.
- *CuidaTEXT*, a text message intervention for family caregiver support has high feasibility, acceptability, and preliminary efficacy, and has potential for widespread implementation.

## Data Availability

All data produced in the present study are available upon reasonable request to the authors

## Acknowledgements

Dr. Perales-Puchalt thanks the national and local organizations that have partnered with him to conduct present and past research since 2015. The research team thanks research participants included in all stages of this research as well as anyone who has contributed directly and indirectly to this research. We also thank the UsAgainstAlzheimer’s A-List and the Alzheimer’s Association’s TrialMatch for sharing the opportunity to participate in *CuidaTEXT* with their registries and website visitors. The ideas and opinions expressed herein are those of the authors alone, and endorsement by the authors’ institutions or the funding agency is not intended and should not be inferred.

---

**Appendix 1.** Ancillary analysis: Preliminary efficacy outcomes comparing pre- and post-intervention scores; including only the first participant from each IWD-cluster.

|  |  | Baseline |  | Follow up |  |  |
| --- | --- | --- | --- | --- | --- | --- |
| **Person with dementia** | N | M | SD | M | SD | P-Value |
| FAQ dependence (0-30) | 17 | 24.9 | 5.1 | 25.9 | 5.4 | 0.386 |
| NPI-Q Severity (0-36) | 17 | 16.6 | 8.4 | 12.1 | 9.5 | **0.010** |
| **Caregiver** | 17 |  |  |  |  |  |
| NPI-Q Distress (0-60) | 17 | 21.0 | 14.2 | 12.2 | 12.8 | **0.001** |
| Caregiver Strain Index (0-26) | 17 | 13.8 | 5.5 | 11.1 | 6.5 | **0.011** |
| ZBI Burden (0-24) | 17 | 10.6 | 5.2 | 9.3 | 6.4 | 0.265 |
| PCS Competence (0-4) | 17 | 2.1 | 0.7 | 2.6 | 0.7 | **0.001** |
| Positive aspects of caregiving (9-45) | 17 | 33.2 | 8.3 | 34.9 | 7.8 | 0.310 |
| Unmet needs (0-34) | 17 | 15.4 | 9.1 | 8.9 | 7.3 | **0.001** |
| EEDS ADRD knowledge (0-14) | 18 | 10.1 | 1.7 | 11.1 | 1.5 | **0.024** |
| ISEL-12 appraisal support (4-16) | 17 | 12.7 | 3.1 | 14.1 | 2.3 | **0.021** |
| ISEL-12 belonging support (4-16) | 17 | 12.2 | 3.4 | 12.6 | 2.9 | 0.624 |
| ISEL-12 tangible support (4-16) | 17 | 12.6 | 2.7 | 13.0 | 3.3 | 0.623 |
| COPE 28 problem focused (8-32) | 17 | 21.5 | 5.9 | 24.5 | 5.1 | 0.074 |
| COPE 28 emotion focused (12-48) | 17 | 31.1 | 5.2 | 32.9 | 6.2 | 0.180 |
| COPE 28 avoidant focused (8-32) | 17 | 13.3 | 3.3 | 12.4 | 3.2 | 0.200 |
| CES-D-10 depression (0-30) | 17 | 8.6 | 4.2 | 5.2 | 4.4 | **0.016** |
| SPANE-Positive affect (6-30) | 17 | 22.1 | 5.1 | 25.1 | 3.5 | **0.003** |
| SPANE-Negative affect (6-30) | 17 | 13.0 | 3.1 | 11.6 | 3.1 | 0.134 |
| Perceived health (1-4) | 17 | 2.4 | 0.9 | 2.4 | 0.7 | 0.999 |

## Notes

Conflict of Interest Disclosures: Each author signed a form for disclosure of potential conflicts of interest. No authors reported any financial or other conflicts of interest in relation to the work described.

Funding: This work was supported by Grants R21 AG065755, K01 MD014177, and P30 AG072973 from the NIH.

### Competing Interest Statement

The authors have declared no competing interest.

### Clinical Trial

NCT04418232

### Funding Statement

This work was supported by Grants R21 AG065755, K01 MD014177, and P30 AG072973 from the NIH.

### Author Declarations

Ethics committee/IRB of the University of Kansas Medical Center gave ethical approval for this work

